# Multichannel tDCS with Advanced Targeting for Major Depressive Disorder: A Tele-Supervised At-Home Pilot Study

**DOI:** 10.1101/2024.03.04.24303508

**Authors:** Giulio Ruffini, Ricardo Salvador, Francesca Castaldo, Thais Baleeiro, Joan A. Camprodon, Mohit Chopra, Davide Cappon, Alvaro Pascual-Leone

**Affiliations:** Neuroelectrics Barcelona, Barcelona, Spain; Deanna and Sidney Wolk Center for Memory Health at Hebrew SeniorLife, Boston, MA, United States; Hinda and Arthur Marcus Institute for Aging Research at Hebrew SeniorLife, Boston, MA, United States; Department of Neurology, Harvard Medical School, Boston, MA, United States; Division of Neuropsychiatry and Neuromodulation, Massachusetts General Hospital, Harvard Medical School. Boston, MA, USA

## Abstract

**Introduction:** Proof-of-principle human studies suggest that transcranial direct current stimulation (tDCS) over the dorsolateral prefrontal cortex (DLPFC) may improve depression severity. This open-label multicenter study tested remotely supervised multichannel tDCS delivered at home in patients (N=35) with major depressive disorder (MDD). The primary aim was to assess the feasibility and safety of our protocol. As an exploratory aim, we evaluated therapeutic efficacy: the primary efficacy measure was the median percent change from baseline to the end of the 4-week post-treatment follow-up period in the observer-rated Montgomery-Asberg Depression Mood Rating Scale (MADRS).

**Methods:** Participants received 37 at-home stimulation sessions (30 minutes each) of specifically designed multichannel tDCS targeting the left DLPFC administered over eight weeks (4 weeks of daily treatments plus 4 weeks of taper), with a follow-up period of 4 weeks following the final stimulation session. The stimulation montage (electrode positions and currents) was optimized by employing computational models of the electric field generated by multichannel tDCS using available structural data from a similar population (group optimization). Conducted entirely remotely, the study employed the MADRS for assessment at baseline, at weeks 4 and 8 during treatment, and at 4-week follow-up visits.

**Results:** 34 patients (85.3% women) with a mean age of 59 years, a diagnosis of MDD according to DSM-5 criteria, and a MADRS score ≥20 at the time of study enrolment completed all study visits. At baseline, the mean time since MDD diagnosis was 24.0 (SD 19.1) months. Concerning compliance, 85% of the participants (n=29) completed the complete course of 37 stimulation sessions at home, while 97% completed at least 36 sessions. No detrimental effects were observed, including suicidal ideation and/or behavior. The study observed a median MADRS score reduction of 64.5% (48.6, 72.4) 4 weeks post-treatment (Hedge’s *g* = −3.1). We observed a response rate (≥ 50% improvement in MADRS scores) of 72.7% (n=24) from baseline to the last visit 4 weeks post-treatment. Secondary measures reflected similar improvements.

**Conclusions:** These results suggest that remotely supervised and supported multichannel home-based tDCS is safe and feasible, and antidepressant efficacy motivates further appropriately controlled clinical studies.

## 1. Introduction

Major depressive disorder (MDD) is a pervasive and debilitating mental health condition that affects millions of individuals worldwide (Kupfer, 2012). The overall point prevalence of depressive disorders in Europe is estimated to be 6% and higher in women (8%) than in men (5%) (Arias, 2021), possibly due to differences in biopsychosocial, psychological, and environmental factors (Kuehner, 2017). The one-year and lifetime prevalence of depression has been estimated to be 10.4% and 20.6%, respectively (Hasin, 2018). Furthermore, recent evidence indicates a rising incidence in youth (Liu, 2020), with MDD-afflicted adolescents up to thirty times more likely to commit suicide (Stringaris, 2017). MDD is characterized by a persistent first-person experience of sadness, hopelessness, lack of interest or pleasure in activities, and associated cognitive, behavioral, and autonomic dysfunction, with 30% of patients with treatment-resistant depression attempting suicide at least once in their lives. Beyond the devastating impact on personal well-being, MDD carries substantial economic costs, including healthcare expenses and reduced work productivity (Adorjan & Falkai, 2019).

About 20–40% of patients do not benefit sufficiently from conventional antidepressant therapies, including trials of medication and psychotherapy (Greden, 2001). Pharmacological treatments have limited efficacy, side effects are common (Carvalho, 2016), and one-third of patients are medication-resistant (Rush, 2006) and experience recurrent depressive episodes (Nemeroff, 2007). For patients with treatment-resistant MDD, several neuromodulation strategies offer potential relief, such as electroconvulsive therapy (ECT) and repetitive transcranial magnetic stimulation (rTMS) (Hyde, 2022). While these treatments are safe and effective, they often come with significant costs, potential side effects, and the need for complex equipment and highly trained staff, making them less accessible in regions lacking specialized facilities. Moreover, device neuromodulation therapies require complex logistics, including daily ambulatory visits over several weeks for TMS or the need for a chaperone to transport patients to and from the ECT service thrice or twice a week, given the use of general anesthesia: these logistical requirements associated with clinic-based treatments continue to impose barriers for access to care with device neurotherapeutics. This accessibility issue is particularly problematic for elderly populations who face additional mobility restrictions and require assistance and support to access outpatient clinic services. Indeed, it is estimated that approximately 15% of the elderly (aged > 65) experience clinically significant depressive symptoms (Blazer, 2003), which can lead to increased morbidity and early mortality (Schulz, 2002). Additionally, older age significantly predicts a more challenging progression of depression (Mitchell & Subramanian, 2005), including a lower likelihood of treatment response (Licht-Strunck, 2007; Tedeschini, 2011), reduced prospects for functional recovery (Little, 1998) and increased risk of relapse (Beekman, 2002). Developing safe and effective home-based neuromodulation therapies can help address access to care and scalability challenges (Gersten, 2022).

In an earlier study (Cappon, 2022), we investigated the feasibility of an innovative protocol where multichannel tDCS is administered at home for older adults with MDD, supported by a caregiver (N=5). This investigation employed a multichannel electric field-informed montage (Ruffini, 2014) and a remotely hosted training program to equip caregivers with the necessary knowledge and skills to administer tDCS at home, eliminating lab visits (Cappon, 2022). Based on this preliminary work, we conducted the present home tDCS pilot study of subject and subject-administrator device utilization, remotely supervised and supported home-based tDCS for antidepressant treatment of adult patients aged 22 and older with MDD who had failed to get satisfactory improvement from at least one prior antidepressant medication in the current episode. This study includes several innovative elements, including advanced electric field-informed montage design methods and multichannel tDCS home technology.

### tDCS

tDCS is a method for noninvasive brain stimulation based on decades-old observations that neuronal firing is modulated by low-amplitude electrical direct current (DC). When applied to the cerebral cortex, cathodal DC suppresses neuronal firing (Creutzfedt, 1962; Nitsche, 2000), while anodal DC increases neuronal firing and leads to increased excitability in the targeted cortex. More precisely, our present understanding indicates that the electric field associated with tDCS currents by Ohm’s law is responsible for the depolarization or hyperpolarization of the soma membrane of elongated neurons (Pyramidal cells) and possibly, others to a lesser extent (Molaee-Ardekani, 2013; Galan, 2023), depending on the direction of the field relative to the orientation of the cells (Ruffini, 2013, 2014, 2018): the electric field component normal (orthogonal) to the cortical surface will depolarize the soma of pyramidal neurons if it is pointing “inward” at that location (from apical dendrite to soma), and vice-versa. With multichannel tDCS, it is possible to choose the position, intensity, and polarity of the electrodes and currents to optimize stimulation at a chosen target map involving one or more regions (a cortical network). Low-intensity, controlled currents (typically ∼1 mA and <4 mA) are applied through scalp electrodes in repeated 20-60 min sessions. The resulting subtle but persistent modulation of neuronal activity is believed to lead to plastic effects derived from Hebbian mechanisms. Notably, tDCS-generated electric fields can interact with functional brain networks (Ruffini, 2018), thus enabling the modulation of neurophysiological dynamics and brain connectivity related to mood disorders and MDD.

A recently emerging technology is model-optimized multichannel tDCS (Ruffini, 2014). This technology relies on using realistic physical models (derived from finite element models created from anatomical MRI) of current flow to estimate the electric field generated by a particular multichannel montage. New systems such as *Starstim* (*Neuroelectrics*) employ up to 32 electrodes with relatively small contact areas of a few square centimeters to precisely control the electric field delivered to the cortex. If a cortical stimulation scheme is prescribed by a clinician or derived from physiological brain models (Ruffini, 2018), this technology allows to configure electrode currents to target the desired area.

Hundreds of trials have demonstrated that when appropriate guidelines are followed, tDCS is easy to use, safe and extremely well tolerated (Antal, 2017) both in the clinic and in remotely supervised home tDCS (Pilloni, 2023; Woodham, 2024).

### tDCS studies in MDD

There has been a fairly large number of studies, including randomized, sham-controlled clinical trials (RCTs) on the effects of tDCS in MDD. Results have been variable and, in part, discrepant. For example, Brunoni et al. (2017) found tDCS to have similar efficacy to antidepressant medications, while Loo et al. (2018) found no efficacy of real tDCS over sham. Nonetheless, several meta-analyses have concluded that tDCS is effective for MDD (Mutz, 2018; Brunoni, 2016). Razza et al. (2020) provided a systematic review of all studies of tDCS for the treatment of acute major depressive episodes completed up to January 2020. They included all randomized, sham-controlled RCTs enrolling participants with an acute depressive episode, a total of 23 RCTs with 1,092 participants. They found that active tDCS was superior to sham regarding endpoint depression scores, response, and remission rates. Moreover, active tDCS was safe with a side-effect profile comparable to sham. Moffa et al. (2020) recently published an individual patient data (IPD) meta-analysis evaluating the efficacy and acceptability of tDCS for the treatment of acute major depressive episodes. Moffa (2020) included data from all published placebo-controlled trials on tDCS as the only intervention in MDD conducted until December 2018. This included 9 eligible studies with a total of 572 participants. They found active tDCS to be significantly superior to sham for an antidepressant response (31% vs. 19% respectively; OR = 1.96), remission (20% vs. 12%, OR = 1.94), and depression improvement (effect size β = 0.31). Moreover, they found a consistent, continuous clinical improvement after the end of the tDCS treatment course. It is noteworthy that the clinical efficacy was substantially higher in the studies where the tDCS course was longer (3-4 weeks versus 1-2 weeks). Zhang (2023) conducted a comprehensive meta-analysis to evaluate the antidepressant efficacy of tDCS as a nonpharmacological treatment for depression. By reviewing randomized controlled trials up to December 30, 2020, the analysis included 27 studies with a total of 1204 patients, comparing 653 patients receiving active tDCS treatment to 551 receiving sham tDCS. The results indicated that active tDCS significantly improved depressive symptoms over sham treatments, with a moderate effect size (g = 0.46). Although active tDCS showed superiority in increasing response and remission rates, these differences were not statistically significant. Dropout rates between active and sham tDCS groups were similar, suggesting comparable tolerability. The findings suggest that tDCS, particularly with specific parameters such as a 2 mA stimulation current for 30-minute sessions and in patients not on antidepressants, holds promise as a treatment modality for depressive episodes.

The variability in the literature on the antidepressant effects of tDCS may reflect differences in patient selection as well as in the tDCS protocol. Longer courses of treatment seem particularly important to ensure sustained, lasting benefits. Consistent with the current understanding of mechanisms of action, tDCS antidepressant effects may involve long-term neuroplastic changes that take time to develop and may, in fact, continue to evolve and mature even after the tDCS treatment course has ended. This makes long treatment courses with maintenance phases important and home-based interventions appealing. Importantly, across all studies, active tDCS has been well tolerated, and there have been no significant adverse or side effects.

### tDCS at home

As a relatively simple and portable technology, tDCS is particularly well suited for remotely supervised, home-based treatment. Several equipment manufacturers have developed systems for remotely supervised, home-based use, where the treatment is administered by the patient or an administrator. Treatment parameters, scheduling, and use can be monitored remotely by clinic or research staff. To date, this has been piloted for the treatment of a number of conditions, including neuropathic pain (Garcia-Larrea, 2019), auditory hallucinations in schizophrenia (Andrade, 2013), attention-deficit/hyperactivity disorder (Leffa et al. 2022), multiple sclerosis (Charvet, 2017; Charvet, 2018; Kasschau, 2015, 2016), Mal de Debarquement Syndrome (Cha, 2016), Parkinson’s disease (Agarwal, 2018; Dobbs, 2018), trigeminal neuralgia (Hagenacker, 2014), vascular dementia (André, 2016), Prader-Willi syndrome (Azevedo, 2017), and, recently, MDD (Charvet, 2023; Woodham, 2024) with promising results.

Palm et al. (2018) completed a systematic review of all available evidence on home use of tDCS until May 2017. They identified 22 original research papers, trial protocols, or trial registrations involving home-use tDCS, mostly as an add-on intervention to cognitive or physiotherapeutic intervention. They showed that treatment adherence was high and side effects minimal, and thus, they concluded that remotely controlled and supervised home-used tDCS was feasible and promising. The experience with home-use tDCS has continued to grow since then.

In the setting of depression, Clayton et al. (2018) reported a case of one patient with comorbid multiple sclerosis and recurrent depressive episodes who received a course of remotely supervised tDCS following ECT treatment. Fatigue and mood ratings improved. More recently, Alonzo et al. (2019) completed a proof-of-principle, open-label trial in 34 participants suffering from MDD who were taught to self-administer 20– 28 tDCS sessions (2 mA, 30 min, F3-anode and F8-cathode montage according to 10–20 EEG placement) over 4 weeks followed by a taper phase of 4 sessions 1 week apart. Participants were initially monitored via video link for a few days and then through the completion of an online treatment diary. One participant withdrew from the study due to too many missed sessions. The remaining 33 participants completed 93% of the scheduled sessions in the initial 4-week phase. Ten of the thirteen participants who qualified for the maintenance phase opted to continue. Mood improved significantly from baseline (mean of 27.5 on MADRS) to 1 month after the end of acute treatment (MADRS 15.5; p < 0.001). Side effects reported across 1,149 sessions were minimal, primarily mild to moderate tingling or burning/heat sensation during stimulation and redness at the electrode sites.

Recently (Cappon, 2022), we investigated the feasibility of a protocol similar to the one used in the present study, with multichannel tDCS administered within the homes of older adults with MDD with the help of a study companion (i.e., caregiver). The study, designed by us during the COVID crisis, explored the feasibility of a remotely-hosted training program to avoid visiting the lab. We employed a newly developed multi-channel tDCS system and protocol with real-time monitoring designed to guarantee the safety and efficacy of home-based tDCS. We found that the home-based, remotely-supervised, study companion administered, multi-channel tDCS protocol for older adults with MDD was feasible and safe, paving the way for the design of the larger study described here.

In the study by Charvet (2023), home tDCS was evaluated as a novel therapeutic approach for MDD through an observational clinical trial. This trial involved 16 participants with moderate-to-severe major depressive episodes who underwent 28 sessions of left anodal DLPFC using a bipolar tDCS montage (using 25 cm^2^ sponges on F3/F4) over six weeks, followed by a tapering phase of weekly sessions for an additional four weeks. There were no serious or treatment-limiting adverse events caused by the tDCS intervention, and no participant experienced an increase in depression or suicidality that warranted treatment discontinuation or additional intervention. The findings revealed a significant reduction in depressive symptoms as early as week 2, with continuous improvement noted at each subsequent biweekly assessment. By the end of the acute intervention, responder and remission rates were 75% and 63%, respectively, which increased to 88% and 81% following the tapering period.

In a recent study by Woodham (2024), tDCS (using large rubber electrodes with sponges (23 cm^2^) with anode over F3 and cathode over F4 in the 10/20 EEG system) was evaluated as a home-based treatment for MDD in a fully remote, multisite, double-blind, placebo-controlled, randomized superiority trial conducted in the UK and USA. The study enrolled adults aged 18 years and older who were diagnosed with MDD based on DSM-5 criteria and were experiencing a current depressive episode of at least moderate severity, as indicated by a score of 16 or higher on the 17-item Hamilton Depression Rating Scale (HDRS) but did not have a history of treatment-resistant depression. The study’s protocol included a 10-week blinded phase, consisting of five tDCS sessions per week for the first three weeks, followed by three sessions per week for the subsequent seven weeks. This was followed by a 10-week open-label phase. The tDCS treatment featured 30-minute sessions, where active tDCS was administered at 2 mA and sham tDCS at 0 mA, both with brief ramping up and down phases. A total of 174 participants with MDD were randomized into either the active treatment group (n=87; mean age 37.1 ± 11.1 years) or the sham treatment group (n=87; mean age 38.3 ± 10.9 years). The results revealed a significant improvement in the HDRS scores in the active treatment group, with a mean reduction of 9.4 ± 6.25 points, compared to a mean reduction of 7.1 ± 6.10 points in the sham treatment group (95% CI 0.5 to 4.0, p = 0.012). Concerning MADRS ratings, the active tDCS treatment arm showed a significant improvement from baseline to week 10, with a mean improvement of 11.3 ± 8.81 relative to the sham treatment of 7.7 ± 8.47 (p= 0.006). The effects were evident at week 10, supporting a recent individual patient data analysis, which found that tDCS effect sizes continue to increase up to 10 weeks compared to sham stimulation (Nikolin, 2023). Safety was monitored using real-time assessments through video conference and the availability of a dedicated study number with 24-hour access to researchers. There were no significant differences in the rates of discontinuation between the active (n=13) and sham (n=12) groups. There were no serious adverse events related to the device and no incidents of serious suicide risk.

Using a related technology in a recent study, Gehrman et al. (2024) conducted a fully remote, randomized, controlled trial investigating the efficacy of transcranial current stimulation (using a pulsed waveform instead of direct current) for the acute treatment of MDD. The study compared active and sham treatment among 255 adults who met DSM-5 criteria for MDD and reported moderate to severe depression levels. Over a period of 4 weeks, participants received two daily 20-minute stimulation sessions. The primary outcome was the change in the Beck Depression Inventory-II score, with analyses conducted at 1 and 4 weeks post-treatment. While initial results at week 1 were not statistically significant, the study found a clinically significant improvement in depression symptoms at week 4, particularly in participants with higher adherence rates and notably among female participants.

Finally, some recent studies have produced negative results. In Borrione et al. (2024), a randomized clinical trial assessing the effectiveness of unsupervised home tDCS for major depression, no significant treatment benefits were observed. The study included 210 participants who were administered tDCS with or without a digital psychological intervention versus a sham control. The study protocol involved twenty-one sessions delivered at 2 mA for 30 minutes each day, five days a week for the first three weeks, followed by twice a week for the remaining three weeks. tDCS was administered using large sponge electrodes positioned over the F3 and F4 locations according to the international 10-20 EEG system, with a fixed distance of 10.5 cm from the midline. Participants ensured correct placement of the device with the help of an augmented-reality tool via a smartphone camera. Stimulation was halted if the impedance exceeded 9 kOhm, indicating displacement or removal of the device. For sham stimulation, the setup was identical, but the current was only active for the first and last 45 seconds of each session, peaking at 1 mA. Results indicated no substantial differences in depression severity changes among the groups. Notably, adverse effects such as skin redness and heat sensations were more prevalent in active tDCS groups. In a related study carried out in the clinic, Burkhardt et al. (2023) carried out a multicenter, triple-blind, randomized, sham-controlled study conducted across eight sites in Germany of the efficacy of transcranial direct current stimulation (tDCS) as an adjunct to stable SSRI treatment in adults with MDD was evaluated. Participants aged 18 to 65 who met DSM-5 criteria for MDD and had been on a stable SSRI dose were randomly assigned to receive either active tDCS or sham stimulation. The treatment consisted of 30-minute, 2-mA bifrontal tDCS sessions for 20 consecutive weekdays, followed by two weekly sessions for an additional two weeks. No significant differences were observed in the mean improvement on MADRS after six weeks between the active tDCS and sham groups. The study concluded that tDCS, when used as an add-on treatment to SSRIs, does not demonstrate superiority over sham stimulation in improving depressive symptoms. Mild adverse events were more frequent with active tDCS. The main differences between these negative studies with others discussed above and the study presented in this paper pertain to the montage and current intensity (large bifrontal sponge electrode vs. single target multichannel using small Ag/AgCl electrodes, smaller total injected current) or a reduced number of sessions.

The purpose of the present study was to explore the safety and technical feasibility of a long-duration intervention employing a specifically designed multichannel montage (i.e., electrode locations, current intensity) with the *Starstim* at-home tDCS device in subjects diagnosed with MDD. This pilot aimed at obtaining preliminary data in advance of a larger clinical trial designed to test whether repeated, daily sessions during two months of at-home advanced tDCS can lead to a robust, clinically significant improvement in MDD patients. Our hypothesis is that the use of a more complex but well-designed tDCS montage, together with an increased dose and number of sessions, can lead to higher efficacy and that, despite its increased complexity, this technology is feasible for home use. Finally, our goal was also to explore the duration of effects one month after the end of treatment.

## 2. Methods

### Participants

Inclusion criteria for this prospective, single cohort, multicenter clinical investigation included a diagnosis of MDD according to the Diagnostic and Statistical Manual of Mental Disorders (DSM-5), as determined via a telehealth interview with a study site psychiatrist or study staff physician with experience in the management of MDD, 22 years or older as of the date of study enrolment, experiencing a major depressive episode of at least four weeks duration, and a MADRS score ≥20 at the time of study enrolment without a pre-specified upper or lower limit of failed antidepressant medications in the current episode or lifetime. Participants also had to be taking at least one medication approved by the FDA for the treatment of depression (except bupropion, which can lower the seizure threshold) whose dose had remained unchanged for four weeks before study enrolment. In addition, participants had to identify and designate one or more adults (persons aged 22 or older) as ‘Administrator/s.’ These individuals had to be willing, able, and formally agree to administer the home-based tDCS, be accessible to the study staff, reporting any safety concerns, potential protocol violations, and any other study-related matters. Subjects also needed access to a wireless internet (Wi-Fi) connection where the study treatments were administered. An accurate and current accounting of the study treatments for each subject was maintained on an ongoing basis by the device interface within the NE portal.

Exclusion criteria included any DSM-5-defined psychotic disorder in the three months preceding the date of study enrolment, active suicidal ideation assessed on C-SSRS (Columbia-Suicide Severity Rating Scale, history of clinically defined medically significant neurological disorder, skin lesions on the scalp at the proposed electrode sites, any cranial metal implants (excluding ≦1 mm thick epicranial titanium skull plates and dental fillings) or medical devices (i.e., cardiac pacemaker, deep brain stimulator, medication infusion pump, cochlear implant, vagus nerve stimulator), previous surgeries opening the skull leaving skull defects capable of allowing the insertion of a cylinder with a radius greater or equal to 5 mm. Participants on antidepressant medications (except bupropion) were allowed to enter the trial provided that the medication dose remained unchanged for four weeks prior to enrolment in the trial and there was no planned dose change for the duration of the trial.

The study (<u>NCT05205915</u>, clinicaltrials.gov) was approved by the WCG-IRB (Western Institutional Review Board-Copernicus Group), and written informed consent was obtained from each participant before the start of study-specific procedures. Because of the nature of this study, consent was obtained electronically online. Information was provided both verbally and in writing, and subjects (or their legal representatives) had ample opportunity to inquire about the details of the study. The study was conducted according to the Declaration of Helsinki, Protection of Human Volunteers (21 CFR 50), Safety reporting in clinical investigations of medical devices under Regulation (EU) 2017/745, Institutional Review Boards (21 CFR 56), Obligations of Clinical Investigators (21 CFR 812), and Clinical Investigation of Medical Devices for Human Subjects – Good Clinical Practice (ISO 14155:2020). The clinical investigation was approved by the FDA (protocol number: NE-02, version 5 dated January 22nd, 2022 (FDA approval letter RE: G160208/S010 dated March 3, 2022) and WCG-IRB on January 31st, 2022).

Results from other home studies suggested that approximately 30 subjects were appropriate to establish preliminary evidence of the safety, tolerability, and feasibility of home administration. Concerning the exploration of efficacy, robust intervention effects (follow-up vs. baseline) were observed with this sample size in a similar open-label study (Alonzo, 2019). Formal sample size calculation in this open-label study was not applicable. Participants were recruited from five centers in the United States (three in Florida, one in Oklahoma, and one in Georgia, v. <u>NCT05205915</u>, clinicaltrials.gov, for more information).

### Protocol

This study was conducted on a “virtual” basis with patients recruited at four U.S.-based sites selected for their specialized expertise and infrastructure dedicated to the efficient management and execution of clinical trials. All visits were remote. The treatment course (see Figure 1) consisted of an acute phase of 28 tDCS sessions conducted daily (7 days per week) over four weeks, consistent with the protocol of Alonzo et al. (2019) and our prior study (Cappon, 2022). This was motivated by the results of Brunoni et al. (2016) and the meta-analysis of Moffa et al. (2020), which found a positive association between increased tDCS ‘dose’ and treatment efficacy. After that, participants underwent a taper phase of an additional 9 sessions of tDCS applied in progressively decreasing frequency until day #60 of the study as follows: (i) Three tDCS sessions once every other day, (ii) three tDCS sessions once every third day, (iii) three tDCS sessions once every fourth day. An incomplete session was defined as one that discontinued stimulation before 100% completion and could be repeated within 24 hours if less than 75% of the session was delivered to the subject. A missed session (0% stimulation delivered) was defined as an anticipated session that did not occur within 24 hours of the assigned date/time. The Montgomery-Asberg Depression Rating Scale (MADRS) (Montgomery & Asberg, 1979) was completed at baseline, approximately at days #28 (end of acute phase) and #56 (end of taper phase) of treatment, and at the end of the 4-week follow-up period.

**Figure 1.**
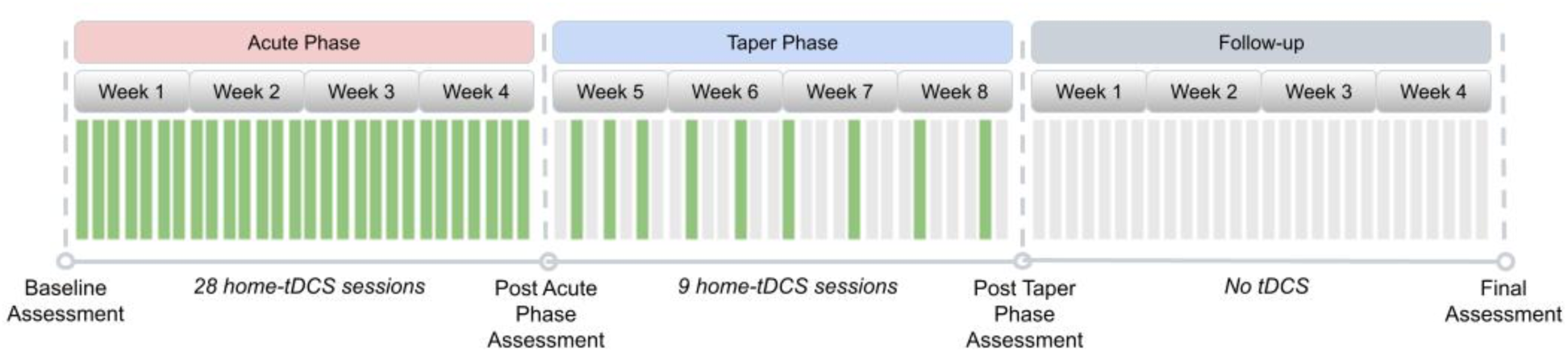
Study Design. The design included an Acute Phase with 28 home tDCS sessions followed by a Taper phase during four weeks. Assessments were all remote. Green bars indicate days with a stimulation session, and grey bars indicate days without a stimulation session. Assessments occurred at four time points – baseline, post-acute treatment, post-taper, and at follow-up four weeks after the end of treatment.

### Multichannel tDCS montage

Stimulation was applied using the *Starstim* device, with current delivered via four *NG Pi* electrodes (circular Ag/AgCl electrodes using gel with a contact area of 3.14 cm^2^) embedded in a neoprene cap. All study subjects used the same fixed montage (electrode locations and currents). The left dorsolateral prefrontal cortex (L-DLPFC) has been consistently related to depression symptomatology (Mayberg, 2001; Pizzagalli, 2011). Specifically, the L-DLPFC is hypoactive in depression, and an increase in activity is associated with antidepressant response. The stimulation target for this study is shown in Figure 2. This target region was selected because it encompasses many clinically validated transcranial magnetic stimulation (TMS) targets for refractory MDD, including those proposed by Fox (2012), Mir-Moghtdaei (2015), Herbsman (2009), Rusjan (2010), and Fitzgerald (2009). Consequently, we designed the multichannel tDCS montage with the maximal normal (orthogonal to the cortex) component of the electrical field targeting the L-DLPFC (excitatory, with the component pointing from CSF into gray matter) with minimal off-target stimulation and for administration via four *NG Pistim* electrodes (3.14 cm^2^ Ag/AgCl gel electrodes) using the *Starstim®-Home* system (see Figure 3 for montage design and the *Starstim Home* system).

**Figure 2:**
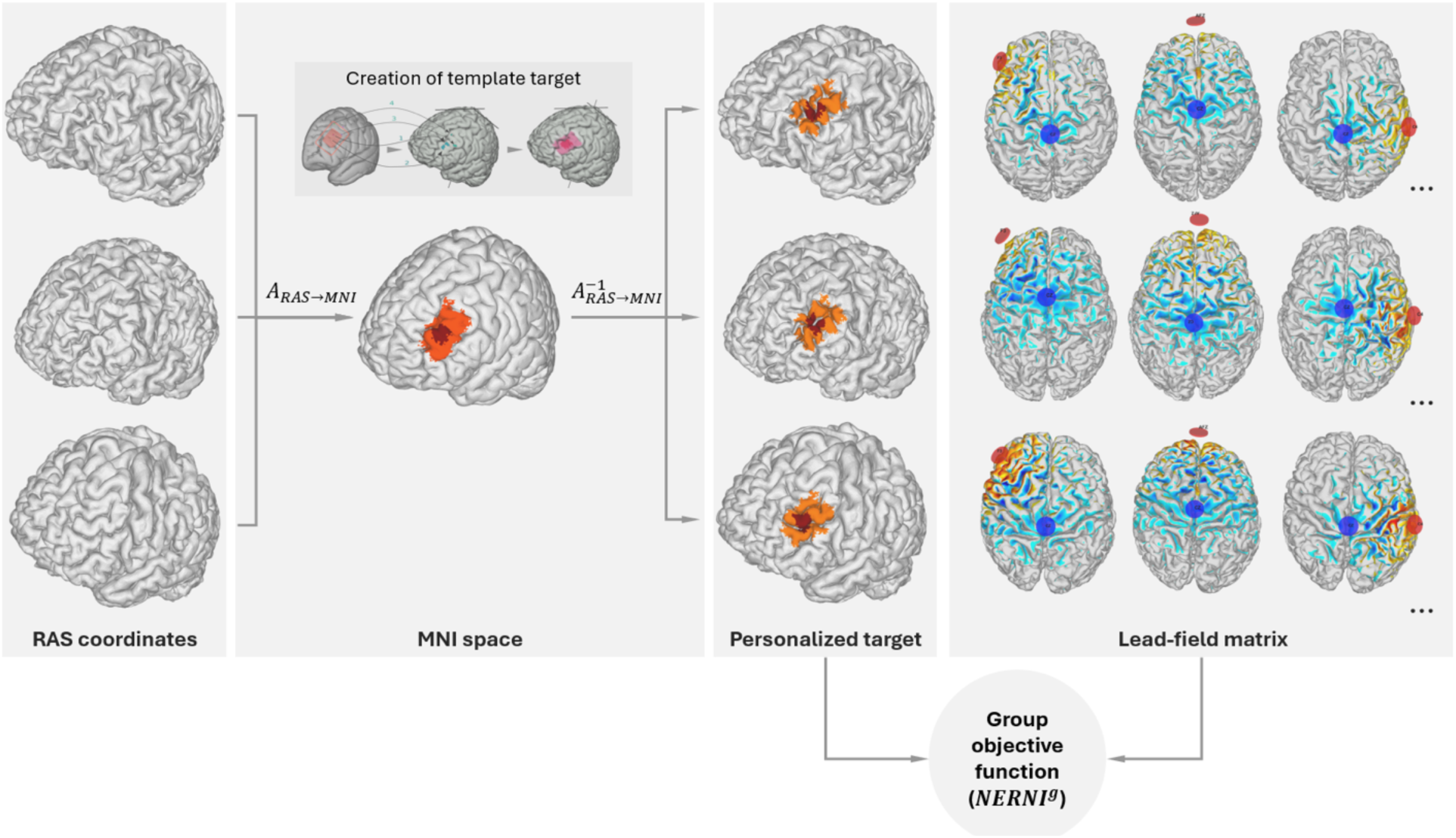
Target definition and mapping to the individualized brain model of each subject. **The central inset (“Creation of template target”)** in the **MNI space** column provides a view of the target specification process. The target consists of a central region (dark red) surrounded by a buffer region of lower weights (in orange). The red rectangle represents the left DLPFC derived from evidence-based TMS targets for depression, as reported by Fox et al. (2012) and the *Beam F3* method (Trapp et al., 2020). The MNI coordinates [*x,y,z*] of the TMS hotspots (1: [−40.6, 41.7, 34.3; −41.5, 41.1, 33.4], 2: [39.3, 46.2 27.5; −41.3, 48.9, 27.7], 3:[−50, 30,36], 4: [−33.6, 30.8, 51.11]) were remapped on the cortex of a default brain model. To obtain the target map in the model, we drew an inner hotspot area encompassing all the mapped points and surrounded it by a buffer area. **Group Objective function creation**: An individualized transformation is derived by mapping the brain model of each subject from RAS (Right, Anterior, Superior) coordinates into MNI (Montreal Neurological Institute) space. The target map in MNI space is then projected into the brain of each of the database subjects using the inverse transformation (from MNI to RAS coordinates), as described in the main text. The group-objective function (*NERNI^g^*, a normalized version of the *ERNI* described in Ruffini et al., 2014) takes as inputs a weighted target map for each of the subjects. The calculation of the objective function also requires the *Lead-field matrix,* which is assembled by calculating all possible bipolar calculations with *Cz* as a common cathode (−1 mA) and the other electrode as an anode (+1 mA), as discussed in Ruffini et al., 2014 (right panel).

**Figure 3.**
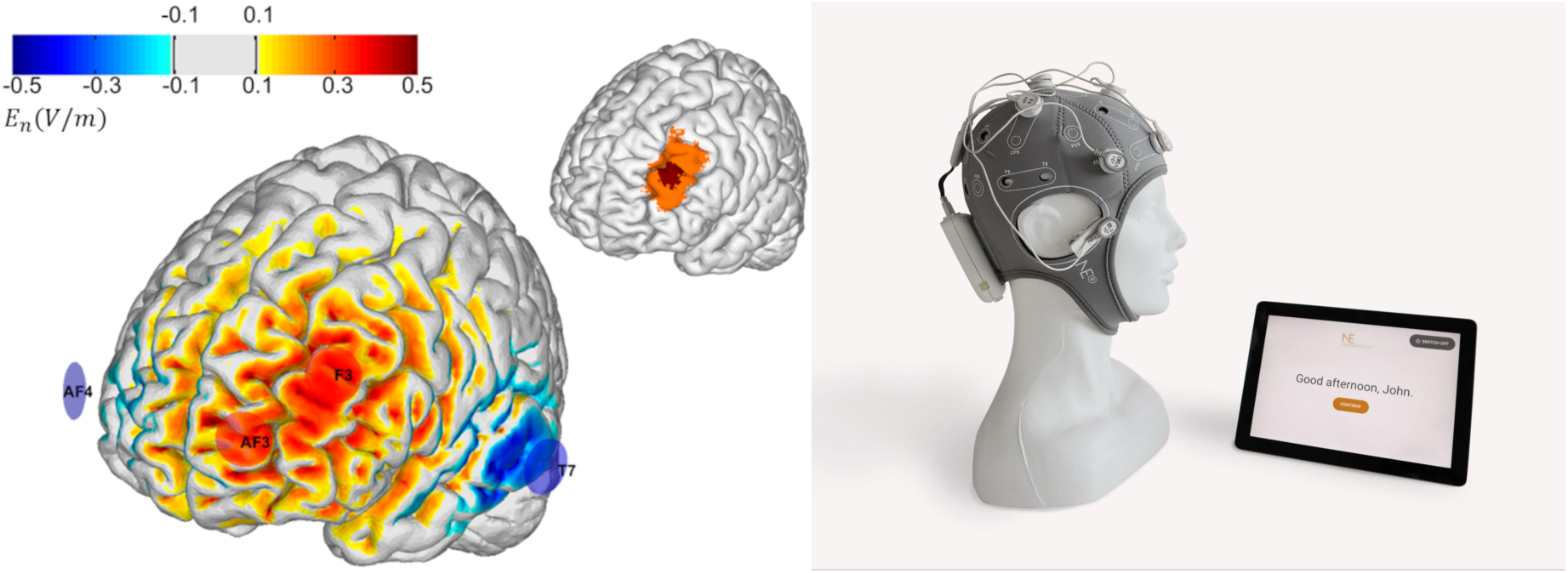
Left: Montage design produced by group optimization. **Left**: The selected group optimized montage consisted of 4 electrodes: two anodes located over the target (AF3 and F3) and cathodes located further away (T7 and AF4). The color scale represents *En*, the normal component (normal to the cortical surface with red/blue denoting inward/outward E-field normal component) of the E-field induced by the optimized montage in the cortical surface (in V/m). **Right:** Starstim Home system (including tablet).

To create a well-targeted but non-personalized montage appropriate for use across many subjects, we used the *Stimweaver®* algorithm (Ruffini, 2014) with *Group Optimization* (GO, Salvador, 2021). The original *Stimweaver®* algorithm explores the space of electrode locations and currents to match the produced electric field with the desired weighted target map, minimizing an Objective Function (OF) that reflects the error of the match for a particular subject. In GO, the objective function is defined as the average OFs of many subjects from an anatomically representative MRI dataset, as shown in Figure 2. In this particular case, we performed a group optimization over 27 healthy patients with an age range between 18 and 93 (55±25 years old). The calculation of the group OF requires the calculation of the lead-field matrix (see Figure 2) for each subject, calculated from personalized biophysical head models created by using the methods summarized in Mercadal et al., (2022). A target map with the weighted target En (the component of the E-field normal to the cortical surface) is also required for each subject. The target map used in this study (left DLPFC) was first defined in the cortical surface of a reference head model in MNI space (Colin27 brain template). This was done by defining MNI coordinates of regions previously defined using several criteria (also described in Cappon et al., 2022) (see Figure 2) from areas identified by neuronavigated TMS, areas activated by working memory tasks identified by fMRI, and areas associated with the subgenual cortex based on rs-fMRI data. These MNI coordinates were clustered into a core area, assigned to higher weights in the optimization algorithm, and surrounded by a buffer area with lower weights. This is shown in Figure 2 (inset box: *Creation of template target*). This target was then mapped to the cortical surface of each of the subjects used in the group optimization, as displayed in Figure 2: each cortical surface was mapped to MNI space using an individualized affine transformation calculated by Freesurfer (v6.0.0, https://surfer.nmr.mgh.harvard.edu/); then, in MNI space, the coordinates of the nodes of the target area defined in the template brain were assigned to the closest node in the personalized cortical surface. The desired En-field in the target region was set to 0.75 V/m (with weights set to 8 for the buffer region and 10 for the core area). The rest of the cortical surface was assigned a 0 V/m target En with a lower weight of 2. The montage was constrained to a maximum of four stimulation electrodes for ease of use by participants at home. The currents were limited to 1.7 mA max per electrode (in absolute value) and 4.0 mA for the total injected current (here defined as the sum of current in all the anodes), well below the recommended safety limits (Antal, 2017). The total injected current in the group-optimized montage was 3.1 mA. The electrode positions found were AF3 and F3 (anodes) as well as T7 and AF4 (cathodes), according to the 10-20 EEG system (Figure 3). The average normal En-field on the target produced by this montage ranged from 0.07 V/m to 0.26 V/m (0.13±0.04 V/m), where positive numbers indicate the field direction pointing into the cortex (with excitatory effects according to the first order model of membrane perturbation of pyramidal cells, Ruffini 2014). In the rest of the non-involved cortex, field amplitude remained low: −0.002±0.001 V/m. For all participants, the current intensity was ramped up over 30 s, then sustained at the stimulation intensity for 30 min, and then ramped down over 30 seconds.

### Home tDCS system

This study used the *Starstim Home Kit* (Neuroelectrics, see Figure 2). Neuroelectrics developed this system for home-based tDCS, effectively overcoming previous challenges with other forms of tDCS and used in several studies, e.g., Garcia-Larrea 2019 (NCT02346396). The *Starstim Home Kit* uses Neuroelectrics’ *Starstim* system with additional features that allow researchers and clinicians to “prescribe” and monitor home-based tDCS to end users. The users could communicate in real time with remote study staff via video-conferencing during device training and during the first three use sessions. The *Starstim* system includes an EEG-like neoprene headcap with holes located where small electrodes can be attached and secured in place in the correct position on the scalp. These electrode holes are color- and number-coded so that electrode leads with corresponding colors in the tDCS device are appropriately attached to the corresponding electrodes, eliminating the potential for accidental mismatching of the electrodes and the leads. The *Starstim®-Home Kit* further incorporates a smart tablet wirelessly connected to the internet.

In more detail, the system includes 1) *Necbox*, the portable wireless tDCS device that applies brain stimulation; 2) Neoprene headcap: electrode positioner on which the relevant electrode positions are marked on the headcap with different colors; 3) Color-coded electrode cables: marked with the same colors as the headcap and with numbers visible on the software interface; 4) *Pistim* (3.14 cm^2^) Ag/AgCl electrodes; 5) Tablet with *HomeApp*: a user interface that guides patients throughout the session and ensures correct delivery of the treatment. 6) Neuroelectrics Portal: a web interface that allows investigators to schedule treatment sessions and monitor compliance in real time.

The tablet allowed the study companions and patient participants to initiate the tDCS sessions, receive specific step-by-step instructions needed to complete the tDCS administration process, and record any side effects via custom-developed questionnaires on the tablet. The table provides simplified instructions and step-by-step touchscreen prompts for the participant. This process has been designed for ease of use, even for individuals who are not computer savvy. The tablet automatically runs an impedance check before and during the delivery of the tDCS current and blocks the stimulation if the electrode impedance reaches above 20 kΩ. Moreover, the tablet has a manual abort function that allows the participant to stop the stimulation if they are experiencing any discomfort or pain. The research staff are notified if this occurs and reach out to the participant to resolve the situation. The tablet further interfaces with another component of the Starstim®-Home Kit called the Neuroelectrics Portal, which the research staff can use to schedule a specific time slot when the execution of the tDCS sessions is allowed. If the stimulation is attempted outside of this time slot, the tablet will inform the participant that the stimulation is currently unavailable and indicate when the next time slot is scheduled. The tablet further allows the study staff to remotely monitor patient participant progression through each session, side effects, and treatment compliance. This portal also ensures that all the stimulation parameters, including stimulation intensity, stimulation duration, and number of sessions, are pre-configured into the system and cannot be adjusted by study companions or patient participants.

Finally, following earlier work described in Cappon et al. (2023), we developed a training and supervision program to accompany the Starstim Home Kit. Study staff members used these training materials to train subjects and administrators on the proposed use of the device. Study staff members monitored treatment sessions until the subject-administrator pairs demonstrated proficiency in all treatment-related procedures, typically through the first three sessions. At the end of each treatment period, the study staff continued to stay in touch with the subject-administrator pairs and inquire about their use of treatment sessions.

### Clinical measures

The main purpose of this study was to obtain preliminary data in advance of a larger clinical trial designed to test whether repeated, daily sessions of at-home transcranial direct current stimulation (tDCS) are feasible and safe and explore if this approach can lead to a clinically significant improvement in patients with MDD.

The Neuroelectrics cloud portal provided information related to electrode impedance, tDCS progress, and tDCS session interruption or termination, whether voluntary or due to a technical issue. These metrics were used to assess feasibility (number of interrupted sessions, missed sessions). Adverse Event collection and concomitant medication evaluation occurred at the start of the acute treatment, start of the taper phase, end of treatment and end of the study, and any Serious Adverse Experiences were evaluated as the primary safety endpoint (SAEs, adverse events occurring at any dose that results in death, a life-threatening adverse experience, inpatient hospitalization or prolongation of existing hospitalization, a persistent, permanent or significant disability/incapacity, required intervention to prevent permanent impairment or damage, a congenital anomaly/birth defect, or other important medical events that may also be considered an SAE when, based on appropriate medical judgment, they jeopardize the study subject or require intervention to prevent one of the outcomes listed).

An exploratory aim of the study was to assess the therapeutic antidepressant efficacy of our protocol. The primary efficacy measure for this study was the median percentage change from baseline to the end of the 4-week post-treatment period in the observer-rated Montgomery-Asberg Depression Mood Rating Scale (MADRS, Montgomery & Asberg, 1979). The secondary outcome measures were: a) Response rate, where clinically significant response was defined as ≥ 50% improvement in MADRS score from baseline to the 4-week follow-up, b) Median percentage change in MADRS score from baseline to the end of week 4 of treatment (acute treatment), to the end of week 8 of treatment (taper phase), c) Change from baseline in the participant-rated Quick Inventory of Depressive Symptomatology (QIDS-SR) (Rush, 2003) administered at the same time points as the MADRS, d) Change from baseline in the Quality of Life Enjoyment and Satisfaction Questionnaire Short Form (Q-LES-Q-SF) (Endicott, 1993), administered at the same time points as the MADRS. Finally, a Safety secondary endpoint was the Change from baseline C-SSRS responses ideation and attempt at any time during acute treatment. C-SSRS evaluation was carried out at contacts between investigator and subject daily during the first 4 weeks of daily stimulation sessions (unless subject discontinues protocol during that time).

### Statistical analysis

This open-label pilot feasibility telemedicine study involved a total of 37 at-home stimulation sessions (30 minutes each) of multichannel excitatory tDCS targeting the L-DLPFC administered over eight weeks, with a follow-up period of 4 weeks following the final stimulation session.

No inferential statistical analysis was planned. The following populations of descriptive analysis were used: a) Safety population (SAF): all participants who have undergone transcranial direct current stimulation at least once (including incomplete stimulation sessions); b) Intention-to-treat (ITT): all participants who have signed the Informed Consent form; c) Per protocol (PP): all participants who have completed at least 75% of the 37 tDCS sessions, have had the final MADRS score recorded and have no major protocol deviations.

For the primary efficacy analysis, the efficacy measure was the median percentage change (MPC) from baseline to the end of the 4-week post-treatment period in the observer-rated MADRS scores. A descriptive analysis of the MADRS at each visit, baseline, week 4, week 8, and at the 4-week post-treatment visit, is also presented. This analysis was performed for both the ITT and the PP sets.

**Figure 3.**
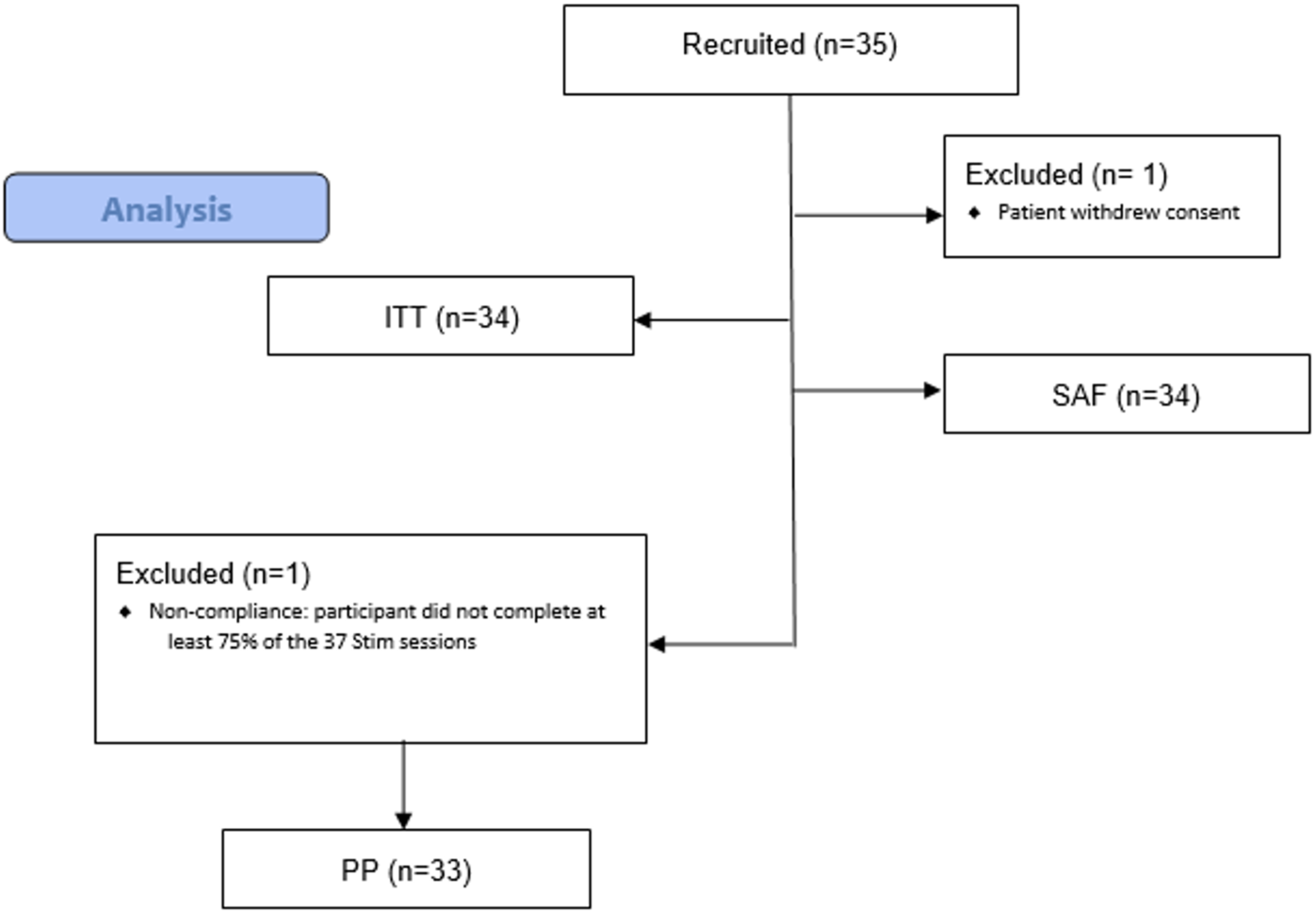
CONSORT flow diagram.

## 3. Results

### Participants

The total valid sample included 35 patients. Figure 3 shows a flowchart of patients recruited and the number and reasons for the exclusion of each population during the study.

At baseline, the study ITT population participants (n=34) were aged between 24 and 78 years, with a mean (standard deviation) of 58.9 (12.9) years. They were primarily female (85.3%). Twenty-one participants (61.8%) were of Hispanic or Latino ethnicity. The mean (standard deviation) time since MDD diagnosis was 24.2 (19.1) months. Additional demographic and education characteristics at baseline for the ITT population are summarized in Table 1.

**Table 1.** Baseline demographic data (ITT)

|  | ITT population<br>(n=34) |
| --- | --- |
| *Variable* |  |
| Age, mean (SD), y | 58.9 (12.9) |
| **Sex at birth** |  |
| Female, n (%) | 29 (85.3%) |
| Male, n (%) | 5 (14.7%) |
| **Ethnicity** |  |
| Hispanic or Latino, n (%) | 21 (61.8%) |
| Not Hispanic or Latino, n (%) | 13 (38.2%) |
| **Race** |  |
| American Indian or Alaska native, n (%) | 1 (2.9%) |
| Black or African American, n (%) | 3 (8.8%) |
| White, n (%) | 30 (88.2%) |
| Head Circumference, mean (SD), (cm) | 56.0 (1.6) |
| **Education level** |  |
| High School Diploma or GED, n (%) | 17 (50.0%) |
| Bachelor's Degree, n (%) | 8 (23.5%) |
| Some college, no degree, n (%) | 5 (14.7%) |
| Did not graduate High School or obtain a GED, n (%) | 2 (5.9%) |
| Academic Associate Degree, n (%) | 1 (2.9%) |
| Master's Degree, n (%) | 1 (2.9%) |

Regarding concomitant psychiatric medications, more than one-third of the patients (12, or 35.3%) were on Sertraline, six (17.6%) were on Citalopram. Three patients (8.8%) were on Duloxetine, three (8.8%) on Memantine, 3 (8.8%) on Quetiapine, and three (8.8%) on Trazodone.

### Safety and Adverse Event Monitoring

Concerning safety, no detrimental effects were observed for the patients. Noteworthy, as measured with the C-SSRS, no participants had suicidal ideation and/or behavior, whether at baseline during treatment or at four weeks post-treatment.

Protocol deviations were evaluated for any trends or patterns that would require additional corrective actions or submissions. All of them were minor, and none resulted in an adverse event or required patient discontinuation from the study. Only 5 (15%) patients experienced adverse events during the study. None of them were reported as serious. Two unexpected adverse events were reported in one patient (3%), and eight adverse device events were reported in four patients (12%). Likewise, no serious adverse device events were reported.

### Feasibility and compliance

85% of the patients (n=29) in the ITT group (n=34) completed all 37 stimulation sessions at home during the acute and taper phases, and 97% (n=33) completed at least 36 sessions (one subject was excluded, see Figure 3).

### Efficacy

The mean (SD) difference between the final visit and baseline for the MADRS score was −19.8 (8.6) for both ITT and the PP population datasets. The primary endpoint (median percentage change in the MADRS score) was 64.5% (48.6%, 72.4%) in both populations.

In assessing the effect size between baseline and week 12 conditions, the pooled standard deviation of MADRS scores was calculated to be approximately 5.8. Cohen’s *d* was 3.1, suggesting a large and statistically significant difference between the group means (to account for the small sample size bias, Hedges’ *g* was also computed, resulting in a value of approximately 3.1). On the other hand, Cohen’s *d*_z_ was 2.0. These statistics reflect the pronounced difference between the baseline and final-visit conditions under study.

The response rate analysis showed that in 73% of patients (n=24), an improvement ≥ 50% was observed in the MADRS score from baseline to the last visit (4 weeks post-treatment, see Figure 4). Finally, improvement was observed from baseline to the end of the study (4 weeks post-treatment) for the QIDS-SR and the Q-LES-Q-SF scores. The mean (SD) and the median (IQR) difference between the final visit and baseline for the Q-LES-Q-SF score were 27.9 (13.8) and 26.8 (17.9, 35.7), respectively.

**Figure 4:**
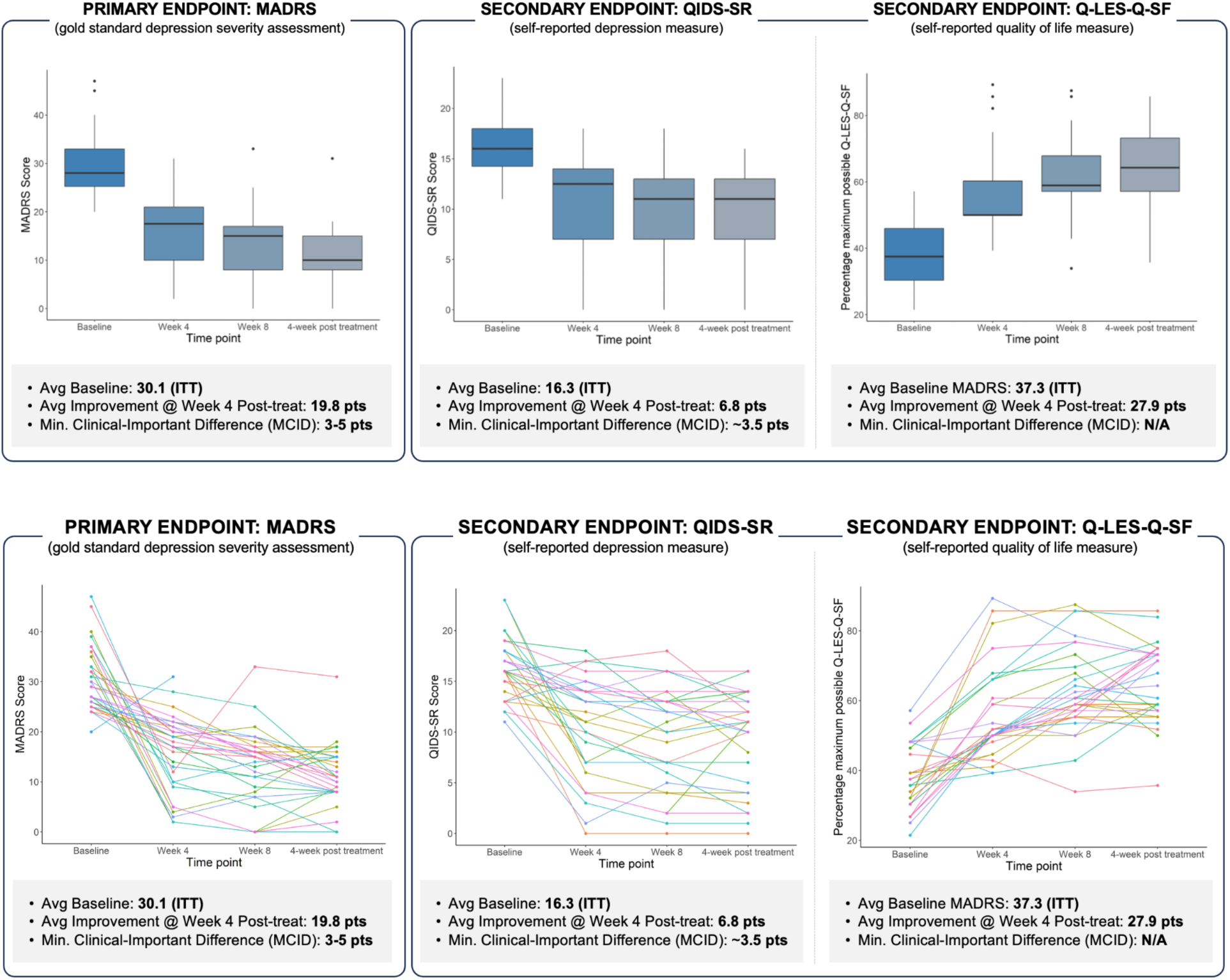
Comprehensive depiction of treatment response over time in patients with Major Depressive Disorder (MDD). Top panel: Boxplots illustrating the distribution of scores (with outliers) at baseline and at weeks 4, 8, and 12. MADRS mean/median scores (STD) at baseline, weeks 4, 8, and 12 post-randomization were 29.8/27 (6.2), 16.7/19 (6.5), 14.4/15 (6.6), and 11.8/11 (5.3). Bottom panel: Longitudinal trajectories of individual patient scores, indicating varied response patterns over the treatment course. The data collectively underscore the heterogeneity in treatment response and the progressive nature of symptom reduction over time.

## 4. Discussion

This exploratory study has demonstrated the feasibility, safety, and potential efficacy of a multichannel home-based, remotely-supervised tDCS intervention with the *Starstim* device in persons with MDD and generated valuable data for planning the next step, i.e., a randomized, sham-controlled, more extensive clinical trial. A single-arm prospective multicenter study with 35 MDD patients was carried out. The population who completed all study visits consisted of 34 patients (85% women and 15% men) with a mean age of 60 years and a MADRS score ≥20 at the time of study enrolment. One patient did not complete at least 75% of all the stimulation sessions. The sample was a representative subset of the MDD population and reflects some of the characteristics of a larger group that could benefit from home-based tDCS.

Regarding primary feasibility objectives, for feasibility, 85% of the subjects completed all the programmed home stimulation sessions throughout the acute and taper phases, and 97% (n=33) completed at least 36 (out of 37) sessions. These positive results confirm the feasibility of the *Starstim* home device and provide crucial information that should be considered for further pivotal studies.

Concerning safety, no detrimental effects were observed for the patients, and all adverse events were minor (see Table 2). Noteworthy, as measured with the C-SSRS, no participants had suicidal ideation and/or behavior, whether at baseline during treatment or at four weeks post-treatment.

**Table 2.** Summary of Mild Adverse Events. No Serious Adverse Experiences were reported, and all Adverse Events were Mild.

| Mild Adverse Events |  |  |  | Relationship to the study device |  |  |  |
| --- | --- | --- | --- | --- | --- | --- | --- |
|  | N° pat.<br>(%) | N° AE | Duration<br>(days)* | Definitely | Probably | Possibly | Unrelated |
| **Total (n=34)** | 5<br>(14.7%) | 9 | 10.3<br>(20.4) | 5 | 2 | 1 | 1 |
| **Skin and subcutaneous tissue disorders** | 3<br>(8.8%) | 4 | 21.7<br>(33.2) | 3 | 1 |  |  |
| Erythema | 1<br>(2.9%) | 1 | 4.0 (.) |  | 1 |  |  |
| Paraesthesia | 1<br>(2.9%) | 2 | 1.0 (.) | 2 |  |  |  |
| Skin burning sensation | 1<br>(2.9%) | 1 | 60.0 (.) | 1 |  |  |  |
| **Nervous system disorders** | 2<br>(5.9%) | 2 | 1.5 ( 0.7) |  | 1 | 1 |  |
| Headache | 2<br>(5.9%) | 2 | 1.5 ( 0.7) |  | 1 | 1 |  |
| **Infections and infestations** | 1<br>(2.9%) | 1 | 11.0 (.) |  |  |  | 1 |
| Sinusitis | 1<br>(2.9%) | 1 | 11.0 (.) |  |  |  | 1 |
| **Musculoskeletal and connective tissue disorders** | 1<br>(2.9%) | 2 | 1.5 ( 0.7) | 2 |  |  |  |
| Myalgia | 1<br>(2.9%) | 2 | 1.5 ( 0.7) | 2 |  |  |  |

The treatment effects were evident at the end of the acute and taper phases and robust four weeks after treatment. The median percentage reduction of the MADRS score was 64.5% (48.6, 72.4), and the mean (SD) difference between the final visit and baseline for the MADRS score was −19.8 (8.6) for both the ITT and the PP population datasets. These results are comparable or superior to those in earlier studies (Wang et al., 2019), as well as the results in Woodham (2024), where the active tDCS treatment arm showed a significant improvement from baseline to week 10, with a change of the MADRS mean score of −11.3 ± 8.8 relative to sham treatment (−7.7 ± 8.5). The results in this study are similar to those in the active arm in the recent placebo-controlled study by Salehinejad et al. (2024). They contrast with earlier recent studies that failed to show efficacy with respect to sham (Burkhardt, 2023, Borrione, 2024). An important difference in our study is the dose and the use of a specifically designed multichannel montage to target the region of interest (these other studies use a standard bifrontal montage with two large sponge electrodes).

Likewise, concerning secondary objectives, in more than 70% of patients (n=24), an improvement of ≥ 50% was observed in the MADRS score from baseline to the last visit (4 weeks post-treatment). The calculated response rate (RR) was 73%. The remission rate in the PP group, evaluated as the percent of participants with a MADRS score equal to or below 10 at the end of acute treatment, taper phase, and four-week follow-up time points, were 30%, 30%, and 52%, respectively. Along the same lines, improvement was observed from baseline to the end of the study (4 weeks post-treatment) for the QIDS-SR and the Q-LES-Q-SF scores. The mean (SD) and the median (IQR) difference between the final visit and baseline for the Q-LES-Q-SF score were 27.9 (13.8) and 26.8 (17.9, 35.7), respectively.

Protocol deviations were evaluated for any trends or patterns that would require additional corrective actions or submissions. All of them were minor, and none resulted in an adverse event or required patient discontinuation from the study.

Considering the good performance of the home-based device plus the overall improvement in depression rating scales (MADRS), symptomatology, and satisfaction questionnaires, it can be said that the developed solution deployed using the Starstim home system was well-accepted and useful for the patients and that it presumably fulfills an unmet need. The Starstim portable multichannel technology proved relatively simple to use and exhibited outstanding performance with a good safety profile. Pending larger controlled trials, this study provides early substantial evidence that home-based, remotely supervised, and supported tDCS treatment with model-designed multichannel montages is feasible for depressed patients and offers a potentially effective intervention. The improved targeting and larger injected current (up to 4 mA) afforded by multichannel *Starstim Home* technology employing multiple electrodes, coupled with its ease of use for repeated, safe stimulation at home, has the potential to deliver more effective solutions. Therefore, this tool may play a significant and outstanding role in applying knowledge to improve the health and healthcare of MDD patients.

### Limitations

Probably the most important shortcoming of this study is the absence of a sham treatment arm. As the effect-sizes to inert “placebo” treatments have gained prominence for psychiatric conditions, especially MDD, the importance of a control condition cannot be underscored enough. The purpose of this investigation, however, was to examine the feasibility of tDCS delivered entirely at home using the *Starstim* portable device with supervision provided remotely, and the study demonstrated that not only was it possible for users to self-administer the intervention but to also derive benefit with improvement in symptoms of major depression. The lack of a control group makes it difficult to argue potential time-dependent changes or to relate changes just to the investigational medical device intervention. However, in comparison with similar studies, the effect size results are very promising. The absence of a control arm also meant that the raters evaluating participants were not blind to the intervention. This was overcome by performing not just objective (MADRS) and subjective (QIDS-SR) assessments of depression severity but also participant reported changes on measures of wellbeing, like the Q-LES-Q-SF. It is also important to note that 21 (of 33 [or 34]) participants were on antidepressant medications and 30 (of 34) participants were on psychotropic medication. Hence, the improvement in MADRS (and other) scores was observed, at least in part, in persons who had been treated for major depression.

The study was conducted remotely and investigators did not assess whether participants had placed the Neuroelectrics Starstim Neoprene cap correctly. While it is possible that some study subjects might not applied the tDCS correctly on the DLPFC, there were a number of safeguards to such errors from happening. The electrode positions on the head-cap and electrode cables were color-coded, and the HomeKit ® tablet provided step-by-step instructions regarding setup, which were specially developed to be simple and easy even for those not familiar with computers. Above all, a web portal allowed study personnel to monitor and assist participants with sessions at any time, allowing proper treatment delivery.

However, this study is one of few of its kind in which a home intervention is being assessed for its impact on MDD well-being. The incremental development of innovative/breakthrough health technologies takes a long time, during which innovation will have to successfully go through testing and evidence generation before it can be launched. As part of this process, early feasibility studies provide the opportunity to capture relevant additional information for the intended use from a real-world setting that would not be possible in non-clinical studies (i.e., bench testing and animal studies) at a very early stage.

Further studies are needed to assess the impact of a digital intervention on MDD, with a longer follow-up period, including a control group and a larger sample size. However, our proof of concept was planned to verify whether the *Starstim* portable technology was feasible and could achieve the desired outcome, and this has been convincingly shown in a real-world setting.

## 5. Conclusions

This pilot study aimed to demonstrate the feasibility of an innovative home-based, remotely supervised, study companion-led, modeling-designed multi-channel tDCS intervention for older adults suffering from MDD in an open-label manner, and available data demonstrates that this was accomplished successfully. The investigation also provided useful safety and preliminary efficacy data for the design of a larger, randomized, controlled at-home trial that will be essential for the broad adoption of tDCS for the treatment of MDD. Since a substantial proportion of patients with major depression show only partial or no improvement after treatment with antidepressants, the availability of additional treatment options would be key to improving the treatment response.

## Data Availability

All data produced in the present study are available upon reasonable request to the authors

## Acknowledgments.

We thank the PIs from the commercial sites in the study for their invaluable contribution to the success of the project and the Clinical Research team at Neuroelectrics for their professionalism and perseverance.

